# Development of a network for interactions and associations among biopsychosocial features of chronic low back pain

**DOI:** 10.64898/2026.02.09.26345929

**Authors:** Pouya Rabiei, Hugo Massé-Alarie, Patrick Desrosiers, the ecobiopsychosocial consortium

## Abstract

**Background:** Understanding the associations among biopsychosocial factors is essential for improving research and treatment of chronic low back pain (CLBP). Here we characterized interrelations among biopsychosocial domains using network analysis and identified the most influential features in CLBP.

**Methods:** Data came from Quebec Low Back Pain Study, comprising 4,489 CLBP participants. We modeled relationships among baseline biopsychosocial features as networks, where nodes represent features and edges encode statistical or causal dependencies among them. Undirected network was inferred using distance correlation. Directed network was constructed using the Linear Non-Gaussian Acyclic Model, which estimates plausible causal directions. Influence maximization was performed using the Independent Cascade (IC) model to identify the most influential features in each network.

**Results:** In the undirected network, physical function and pain interference were the most central nodes, followed by depression. In the directed network, fear of movement, catastrophizing, and widespread pain emerged as key downstream hubs receiving multiple causal inputs, whereas pain interference, physical function, and depression acted as major upstream drivers exerting broad causal influence. IC diffusion simulations further identified pain interference and physical function as the most influential features in the undirected and directed networks, respectively.

**Conclusions:** Pain interference, physical function, and depression consistently emerged as key components of the CLBP biopsychosocial network. These features exert causal effects on fear of movement, catastrophizing, and widespread pain, with diffusion analyses confirming their roles as system-wide drivers. Interventions targeting functionality and pain interference, rather than pain intensity alone, may yield broader benefits across psychological and functional domains.

## 1. Introduction

Network analysis offers a powerful framework to visualize and characterize complex interactions among different indicator. As a broadly interdisciplinary field, ^1^ network science enables the examination of system-level interactions among features across diverse areas like pain science, neuroscience, and sociopsychological science, and provides tools to investigate how they are interrelated ^2,3^. In the field of pain research, low back pain represents a leading cause of disability worldwide,^4^ with approximately 40% of individuals developing persistent symptoms lasting longer than three months, referred to as chronic low back pain (CLBP) ^5^. Growing evidence highlights a range of biopsychosocial factors such as functional impairment, depressive symptoms, maladaptive coping, fear avoidance, stressful life events, and low socioeconomic status that are associated with chronic pain and that may interact to sustain or exacerbate its persistence ^6-9^.However, the nature and organization of these interactions in CLBP remains poorly understood. Network analysis offers a principled approach to uncover these interconnections and to identify key indicator(s) through, for instance, centrality metrics, which quantify the relative importance or influence of each variable within the network ^1,2^.

Lately, the network structure of depressive symptoms in individuals with chronic pain was investigated ^10^. Researchers identified difficulty concentrating, anhedonia (loss of interest or pleasure), depressed mood, and fatigue as the most central features based on network centrality metrics. They suggested that mindfulness-based therapy, which cultivates sustained and nonjudgmental attention to present experience, may reduce vigilance to pain and intrusive thoughts, thereby improving concentration and mitigating depressive symptoms.^10^. A recent study analyzed a large chronic pain cohort (> 10,000), grouping participants by pain extent (number of painful anatomical location). Their undirected network— where the connections between nodes are undirectional— revealed that pain interference was more central in a group of participants with smaller pain extent, while catastrophizing had the highest strength in a group of participants with larger pain extent ^11^.

Most previous pain-related network studies ^12 10,13,14^ have relied on correlation-based association networks, typically using Pearson correlations or regularized partial correlations estimated under Gaussian assumptions (e.g., EBICglasso). While these approaches characterize patterns of co-variation among symptoms, they yield undirected networks and therefore do not provide information about directionality, such as whether one indicator is more likely to influence or precede another rather than merely co-occur. To address these limitations, the present study aimed to construct a comprehensive network of biopsychosocial features associated with chronic low back pain (CLBP) and to identify the most influential factors using data from the Quebec Low Back Pain Study (QLBPS) ^15^.

We adopted a multi-step analytical strategy. First, we implemented two complementary undirected model—distance correlation (dCor)—to quantify pairwise associations under different statistical assumptions. Model of dCor captures general statistical dependence, including potentially nonlinear relationships, without requiring Gaussian assumptions. The most suitable undirected model was selected following the benchmarking framework proposed by Liu et al ^16^. Second, to move beyond association and infer plausible directional relationships, we applied the Linear Non-Gaussian Acyclic Model (LiNGAM)^17,18^ to construct directed networks and identify potential causal hubs within the biopsychosocial system. Together, this combined undirected– directed framework allowed us to characterize both the dependency structure and the directional organization of key indicator(s) underlying CLBP.

## 2. Methods

### 2.1 Study design and participants

This study utilized data from the QLBPS, a longitudinal prospective cohort initiated in November 2018 (clinicaltrials.gov: NCT04791891) ^19-21^. QLBPS is an online-based study that collects data across demographic, pain-related, general health, psychosocial, and occupational domains from individuals who self-identified as having either acute LBP or CLBP. The study has been approved by the Research Ethics Committee of McGill University (A06-M22-18A) and all participants have provided informed consent. The detailed methodology can be found in the QLBPS protocol published previously^15^. The current study only used baseline data from participants recruited from February 2019 to June 2024. This report follows the Strengthening the Reporting of Observational Studies in Epidemiology (STROBE) guidelines^22^.

The QLBPS has recruited participants across the province of Quebec, Canada, through advertisements via social media, newspaper ads, and leaflets in clinical practices. Eligibility criteria include being at least 18 years old, fluency in English or French, and self-reporting LBP in the past 4 weeks ^23^. For the current study, we used data from participants identified as having CLBP within the QLBPS. CLBP was defined based on NIH criteria of CLBP (i.e. pain > 3 months and at least 50% of days in the last 6 months) ^24^.

For the present study, classification of CLBP was based on responses to 2 screening questions in the Canadian adaptation of the NIH minimum dataset regarding LBP duration and frequency, i.e., “how long has low back pain been an ongoing problem for you?” (Less than a month, 1-2 months, 3-5 months, 6-11 months, 1-5 years, or more than 5 years) and “how often has low back pain been an ongoing problem for you over the past 6 months?” (Every day or nearly every day in the past 6 months, at least half the days in the past 6 months, less than half the days in the past 6 months).

### 2.2 Data collection

In the registration phase, potential participants received an invitation to a web platform (backpainconsortium.ca/ or aliases mybackhurts.ca or malaudos.ca). After registration, only those who provided all the required information and self-identified with LBP received an email inviting them to access the baseline online survey, including the informed consent form. Research Electronic Data Capture (REDCap) ^25^ was used to collect data and control the study workflow.

### 2.3 Measures

Baseline data was collected using the Canadian adaptation of the NIH minimum dataset for CLBP ^21,24,26^. This dataset includes 40 items covering a wide range of domains: pain characteristics, comorbid conditions, pain interference, workplace issues related to LBP, physical function, depression, sleep disturbance, fear of movement and catastrophizing, lawsuits, substance abuse, quality of life, socio-demographic data and habits related to smoking and obesity ^21,27^. Domains of the Canadian adaptation of the NIH minimum dataset for CLBP research showed good internal consistency (alpha: 0.81-0.93) ^21,27^.

### 2.4 Network analysis

Network analysis was employed to capture the interactions and associations among a set of biopsychosocial features. These were extracted from Canadian adaptation of the NIH minimum dataset and included socio-demographic (age, sex, employment status, educational level), health status/lifestyle behaviors (body mass index (BMI), smoking habit, quality of life, sleep disturbance, comorbidity) pain-related (pain intensity, pain duration, pain impact, pain interference, widespread pain, neuropathic pain, physical function), and psychological (depression, fear of movement, catastrophizing). Features were selected based on prior evidence linking them to chronic pain and long-term disability ^6,28,29^. All analyses have been done in Python.

Via *sklearn*.*impute* module in Scikit-learn, missing data were managed using the most frequent value strategy in *for* imputation of missing data in categorical features (sex, employment status, smoking habit, pain duration, widespread pain, fear of movement, and catastrophizing), and multiple imputations using chained equations method with 10 imputation rounds for numerical features (age, BMI, educational level, pain intensity, pain duration, pain interference, neuropathic pain, physical function, quality of life, comorbidity, depression, sleep disturbance) followed by z-score standardization.

For the undirected networks, we constructed weighted association graphs from baseline biopsychosocial features. Each indicator was represented as a node, and edges were defined between all pairs of nodes, with edge weights corresponding to the estimated statistical dependence between the associated features. Pairwise statistical dependence was quantified using two measures: mutual information (MI) and distance correlation (dCor), both of which capture linear and nonlinear relationships in heterogeneous data ^30^. For each measure, we computed a pairwise dependence matrix across all features, which was used as a weighted adjacency matrix to construct an undirected graph in Python. Network construction and analysis were performed using NetworkX (version 3.6) ^31^. The primary undirected network was based on dCor. A second network based on MI was constructed for sensitivity analyses. Node centrality was quantified using strength, defined as the sum of the weights of all edges incident to a node ^32^. Higher strength indicates stronger overall statistical dependence between a given indicator and the rest of the network.

Network accuracy and stability were evaluated following the procedure described by Epskamp et al. ^33^. Edge-weight accuracy was assessed using nonparametric bootstrapping (1,000 resamples) to obtain 95% confidence intervals for each edge weight. Centrality stability was examined using case-dropping subset bootstrapping (“m-out-of-n”), in which random subsets of observations were removed, and centrality indices recomputed. Robustness was quantified by computing the correlation between centrality estimates obtained from the full sample and from each subset, providing a measure of the sensitivity of node rankings to sampling variability.

For the directed networks, we constructed directed acyclic graphs (DAGs) from the same set of baseline biopsychosocial features. Each indicator was represented as a node, and directed edges were used to represent putative causal effects from one indicator to another. Directed network inference was performed using LiNGAM implemented in the lingam Python package (version 1.10) ^34^. LiNGAM estimates a directed acyclic graph under a linear structural equation model with non-Gaussian residuals, enabling the identification of plausible causal directions from observational data ^34^. The resulting model yields a weighted adjacency matrix defining a directed graph in which edge direction encodes the inferred causal ordering between features. Centrality in the directed network was quantified using in-degree and out-degree. In-degree quantifies the number of edges directed toward a node, reflecting the extent to which it is influenced by other nodes, whereas out-degree represents the number of edges emanating from a node, indicating its influence on other nodes ^32^.

To evaluate each node’s dynamic influence, we applied influence maximization using the Independent Cascade (IC) diffusion model ^35^. The directed weighted adjacency matrix inferred by LiNGAM was used to define the diffusion network. For each node acting as a single seed, 1,000 simulations were performed with a single seed node selected per simulation, and activation was propagated probabilistically along outgoing edges. At each step, an active node had a single opportunity to activate each inactive neighbor with probability proportional to the corresponding edge weight. Newly activated nodes were added to the active set and allowed to propagate activation in subsequent steps. The diffusion process terminated when no further activations occurred or when a maximum of 50 steps was reached. To obtain stable influence estimates, the diffusion process was repeated for N = 500. Node influence was quantified as the expected number of activated nodes at convergence. Influence maximization was performed by identifying the seed set that maximized the expected cascade size. All diffusion simulations and influence maximization analyses were implemented in Python using the *NetworkX* library.

## 3. Results

4,489 individuals in the dataset who met the criteria of CLBP were included. Table 1 presents the baseline characteristics of the individuals included. eTable 1 in supplementary file shows the labels of the features used for creating the networks and how these are measured in Canadian adaptation of the NIH minimum dataset.

**Table 1.** Baseline characteristics of 4,489 participants with CLBP.

| Baseline characteristics | Mean (SD) or proportion (%) |
| --- | --- |
| Age | 45.92 (12.17) |
| **Sex** |  |
| Male | 1690 (38.39%) |
| Female | 2712 (61.61%) |
| BMI | 29.38 (7.45) |
| Pain intensity<br>(0 – 10) | 6.13 (1.79) |
| **Pain duration (years)** |  |
| 3-5 months | 5.9% |
| 6-11 months | 8.5% |
| 1-5 years | 38.1% |
| > 5 years | 47.3% |
| **Education** |  |
| No diploma | 344 (7.84%) |
| Diploma | 2765 (63.03) |
| Bachelor | 850 (19.38) |
| Master | 359 (8.18) |
| Doctorate | 69 (1.57) |
| **Working condition** |  |
| Fulltime | 2123 (47.29%) |
| Parttime | 456 (10.16) |
| Unemployed, Retired, or<br>Student | 1910 (42.55) |
| **Smoking habit** |  |
| Never smoke | 1884 (43.73%) |
| Current smoker | 852 (19.78%) |
| Used to smoke, but have now quit | 1572 (36.49%) |

BMI, Body mass index; SD, Standard deviation

### 3.1 Features’ collinearity

Before conducting the network analysis, the collinearity has been checked using Pearson correlation. This analysis revealed that the pain impact indicator was strongly correlated (r ≥ 0.80) with pain interference and physical function (Supplementary file, eFigure 1). As pain impact includes four questions of pain interference and physical function and it replicates other two features, we removed it from the analysis to avoid multicollinearity issues.

### 3.2 Undirected network

Inspection of dCor dependence matrix identified the strongest pairwise associations between pain-related and psychological features, in particular among physical function, pain interference, and depressive symptoms. These dominant associations are directly reflected in the corresponding undirected weighted network, where these indicator pairs form the highest-weight edges (Figure 1A). Also, the same pain-related and psychological features showed a densely connected core; however, certain nodes such as sex, BMI, smoking habit, comorbidity, or pain duration peripherally connected, suggesting weaker or more isolated relationships with the rest of the system. Features clustered by domains, with psychological and pain-related forming a tightly integrated subnetwork, suggesting strong statistical interdependence among these features in CLBP, especially between depression and pain-related features.

**Figure 1.**
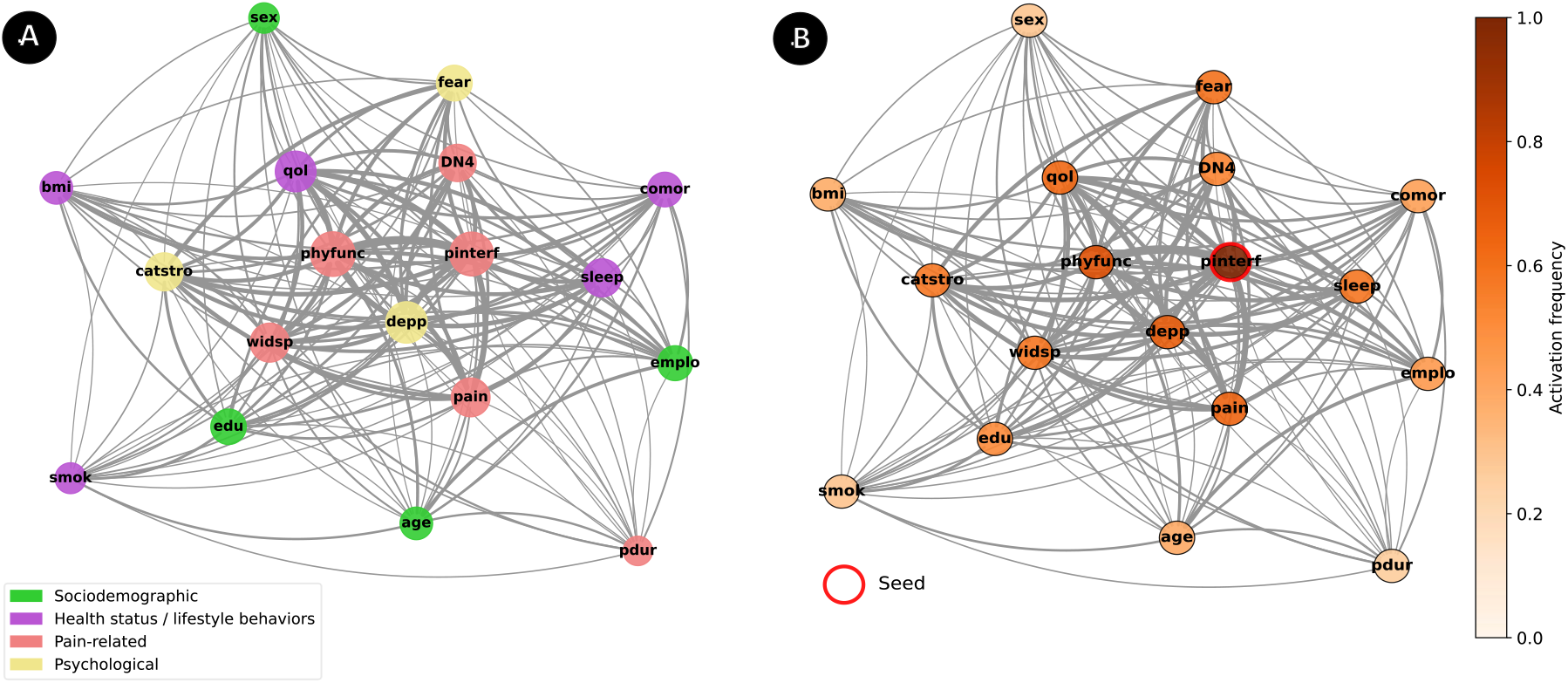
Distance correlation networks of socio-demographic, pain-related, and psychological features. **A:** Undirected network including all features and edges. Nodes represent individual variables (green = sociodemographic, purple = health status/lifestyle behaviors, red = pain-related, yellow = psychological). Edges represented by gray color showed pairwise distance correlations; edge thickness is proportional to correlation strength. Node size reflects node strength (sum of absolute edge weights). **B:** Independent Cascade diffusion in the dCor undirected network. The figure illustrates the simulated diffusion process using the IC model, in which activation initiated from pain interference (highlighted by a red ring) spreads probabilistically through the network according to edge weights and followed by physical function and depressive symptoms. Node color intensity represents the activation frequency with darker shades demonstrating a higher probability of being activated during the diffusion process. Edges are scaled by their connection strength (weight magnitude) derived from dCor coefficients. The simulation was repeated 1,000 times to ensure stability of diffusion estimates, and node colors represent the average activation frequency across runs (darker shades = higher probability of activation). A base activation probability of 0.5 was used, meaning each active node had a 50% chance of activating a connected neighbor, scaled by the strength of their pairwise association (edge weight). Abbreviations: age= age; sex= sex; emplo = employment status; edu= educational level; qol = quality of life; sleep = sleep disturbance; bmi= Body mass index; smok= smoking habit; comor = comorbidity; pain = pain intensity; pdur= pain duration; pinterf = pain interference; widsp = widespread pain; DN4 = neuropathic pain; phyfunc = physical function; fear = fear of movement; catstro = catastrophizing; depp = depressive symptoms

Figure 1B presents the results of the IC diffusion simulation applied to the dCor-based undirected network. Here pain interference is shown as the primary influential nodes in the entire network and then followed by physical function and depressive symptoms. This pattern supports the interpretation of pain interference as a central diffusion hub that can trigger widespread activation across domains within the biopsychosocial network.

Based on the strength centrality (Figure 2), most influential nodes were physical function (strength = 3.87), pain interference (strength = 3.71), and depressive symptoms (strength = 2.89). Other moderately central nodes included quality of life (strength = 2.23), pain intensity (strength = 2.02), and catastrophizing (strength = 1.60).

**Figure 2.**
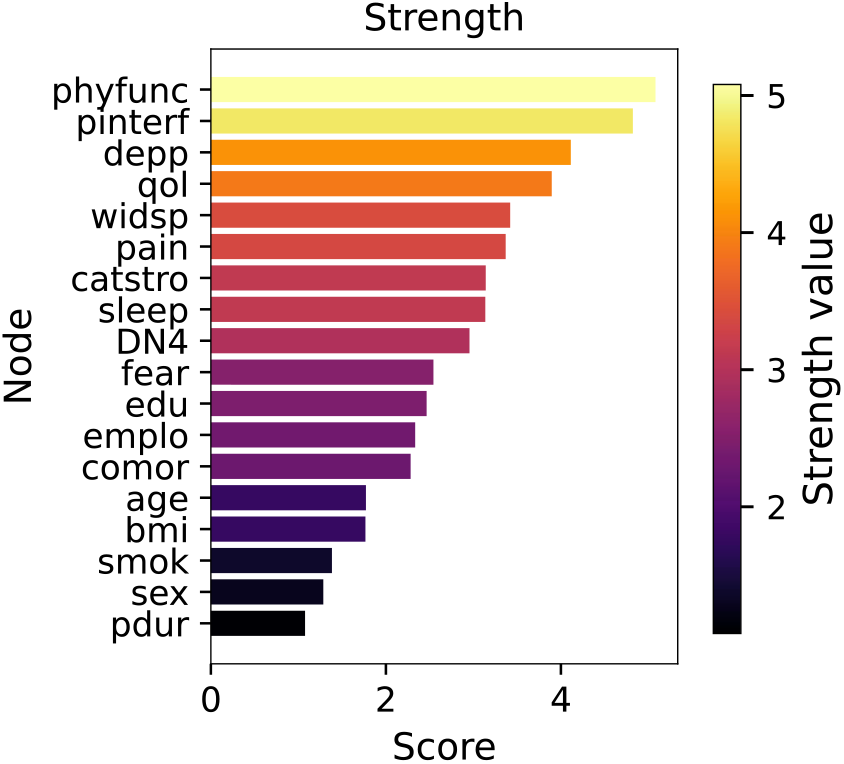
Strength centrality in undirected networks derived from dCor. Bar length and color intensity indicate the normalized strength centrality, representing the sum of absolute edge weights connected to each node. dCor model identified physical function, pain interference, depressive symptoms, and quality of life as nodes with the highest strength centrality, whereas sex, smoking, and pain duration exhibited the lowest values. Abbreviations: age= age; sex= sex; emplo = employment status; edu= educational level; qol = quality of life; sleep = sleep disturbance; bmi= Body mass index; smok= smoking habit; comor = comorbidity; pain = pain intensity; pdur= pain duration; pinterf = pain interference; widsp = widespread pain; DN4 = neuropathic pain; phyfunc = physical function; fear = fear of movement; catstro = catastrophizing; depp = depressive symptoms

Sensitivity analysis also showed that the MI-based network revealed patterns largely consistent with dCor model. In both networks, the strongest and most stable edges connected physical function, pain interference, and depressive symptoms, indicating these features form the central core of the biopsychosocial system (Supplementary file, eFigure 2A). the differences between MI and dCore model was in the influential nodes where unlike dCore, in MI physical disability was the primary influential node (see Supplementary file, eFigure 2B for more details). eFigure 3 in supplementary file summarizes and compares value of strength centrality for all features of two models of dCor and MI. MI model identified the same four features as the most influential nodes (i.e. physical function, pain interference, depressive symptoms and quality of life).

The results of the network accuracy and stability is in the supplementary file, eFigure 4. In brief, bootstrap analyses demonstrated that the dCor network was characterized by a stable and well-defined core structure, with the strongest and most reliable associations centered on pain interference and physical function. Several additional edges involving depressive symptoms, pain intensity, and quality of life also showed robust stability. Centrality stability analyses indicated that strength centrality rankings remained reliable even after substantial case removal, supporting confident interpretation of node importance.

### 3.3 Directed network

The directed network was estimated using Direct LiNGAM, retaining absolute edge weights. Figures 3A and 3B illustrate the resulting directed networks, emphasizing in-degree and out-degree centrality, respectively. In both figures, blue and red edges represent positive and negative causal effects, and edge thickness corresponds to the absolute value of the causal weight, indicating the strength of influence between connected features. In Figure 3A, node size is proportional to in-degree centrality, meaning that larger nodes receive a greater number of causal inputs from other features and are therefore more influenced by other components in the network. Conversely, in Figure 3B, node size reflects out-degree centrality, identifying features that exert stronger causal effects on others and act as potential influencers or drivers within the network.

**Figure 3.**
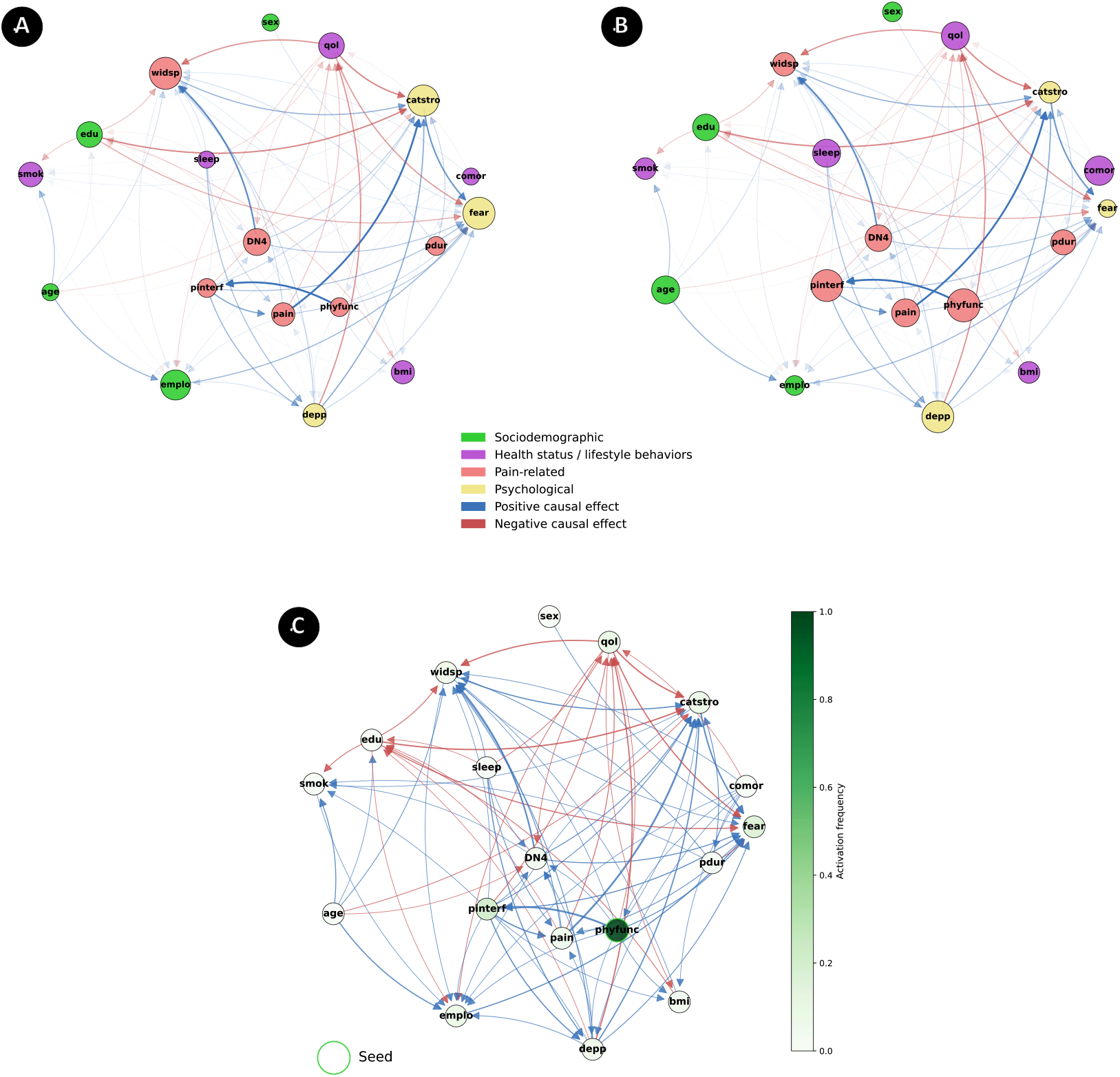
Directed LiNGAM networks and diffusion dynamics of biopsychosocial features. **A:** Directed network estimated emphasizing in-degree centrality. **B**: The same directed network emphasizing out-degree centrality. **C:** Independent cascade diffusion simulation applied to the directed network. For all panels, blue and red arrows represent positive and negative causal effects and edge thickness corresponds to the absolute magnitude of the causal weight, indicating the strength of influence between connected features. For panel A, node size is proportional to in-degree, meaning that larger nodes receive more causal inputs and for panel B node size is proportional to out-degree, meaning that larger nodes send more causal outputs. These figures in above panel show that fear of movement, widespread pain, catastrophizing, and employment status are the major recipients of causal influence while physical function, depressive symptoms, pain interference, and quality of life act as causal drivers. In the below panel, darker green shades denote higher average activation frequency, confirming a consistent and robust propagation pattern from physical function across repeated simulations. This finding reinforces its role as a key driver within the network. Abbreviations: Abbreviations: age= age; sex= sex; emplo = employment status; edu= educational level; qol = quality of life; sleep = sleep disturbance; bmi= Body mass index; smok= smoking habit; comor = comorbidity; pain = pain intensity; pdur= pain duration; pinterf = pain interference; widsp = widespread pain; DN4 = neuropathic pain; phyfunc = physical function; fear = fear of movement; catstro = catastrophizing; depp = depressive symptoms

Accordingly, the strongest positive causal effects were observed from physical function → pain interference (β = 0.45), pain intensity → catastrophizing (β = 0.44), neuropathic pain → widespread pain (β = 0.37), and catastrophizing → fear of movement (β = 0.33). In contrast, the strongest negative causal effects were found from educational level → catastrophizing (β = −0.31), quality of life → catastrophizing (β = −0.31), quality of life → widespread pain (β = −0.28), and depressive symptoms → quality of life (β = −0.28). eTable 2 in supplementary file summarizes all sources and targets according to their weight values of causal effects.

Nodes with the highest in-degree centrality included fear of movement (in-degree = 12), widespread pain (in-degree = 12), catastrophizing (in-degree = 11), and employment status (in-degree = 10). These features act as major recipients of causal inputs within the system. Conversely, nodes with the highest out-degree centrality were physical function (out-degree = 10), depressive symptoms (out-degree = 9), pain interference (out-degree = 9), and comorbidity (out-degree = 7). eTable 3 in supplementary file summarizes all nodes according to their in-degree and out-degree centrality values.

Figure 3B and 3C illustrate the IC diffusion simulation applied to the directed network. Edge color, thickness, and node size follow the same conventions described above, representing the direction, magnitude, and centrality of causal effects, respectively. Diffusion was initiated from physical function, identified as the most influential node within the network. The resulting activation pattern highlights physical function as a central diffusion hub capable of initiating widespread activation across multiple biopsychosocial domains, reinforcing its role as a key driver of system-level interactions.

## 4. Discussion

We aimed to determine the complex associations between biopsychosocial domains using network analysis and determined the most influential features in individuals with CLBP. The network diagram obtained from dCor method showed that pain-related features of pain interference and physical function along with depressive symptoms as a psychological indicator extracted from Canadian adaptation of the NIH Minimum Dataset were the main nodes of centrality metrics. The same pattern was replicated using the MI model, supporting the robustness of these associations. The IC diffusion simulations applied to the dCor network further identified pain interference as a key diffusion hub capable of initiating widespread activation across the system. This finding shows that modifications in pain interference could propagate to other domains, particularly psychological and functional components, reflecting its broad systemic influence.

Our findings from undirected network highlight the key role of pain-related features of, pain interference, and physical function as well as psychological factors such as depressive symptoms. Meanwhile, abovementioned pain-related features were the nodes with the more strength of connection and acted also as bridges/mediators connecting the more peripheral nodes to the central ones. The large strength centrality of pain interference and depressive symptoms was also found in a previous network analysis study of chronic pain individuals ^36^. Moreover, the findings from directed network approach via LiNGAM showed that physical function and depressive symptoms, followed by pain interference emerged as the strongest causal drivers, exerting influence on downstream features such as fear of movement, widespread pain, and catastrophizing. In contrast, fear of movement and catastrophizing—psychological constructs associated to pain avoidance and maladaptive coping—were positively influenced by pain-related features but were negatively impacted by quality of life and educational level. Together, the directed and undirected analyses converge to show that pain interference and physical function play pivotal roles in mediating inter-domain dynamics within the biopsychosocial framework.

Consistent results from influence maximization using IC diffusion on both undirected and directed networks reinforce these observations: pain interference and physical function consistently emerged as dominant nodes whose activation led to the greatest overall propagation. This suggests that interventions targeting improvements in functionality or reductions in pain interference could yield widespread benefits across multiple biopsychosocial domains.

In Canadian adaptation of the NIH Minimum Dataset, pain interference is measured via the questions regarding how pain can affect social and daily activities and physical function also are related to the capability of doing daily activities ^21,27^. Their strong association, confirmed by Pearson correlations in this study, provides a mechanistic basis for their network connectivity. Moreover, prior research has shown that psychological constructs such as depressive symptoms, are highly intertwined with chronic pain and contribute to both current disability and future pain persistence ^6,37-39^ .

Traditional pain assessments focusing solely on pain intensity—such as numeric rating scales— as well as interventions focusing on pain intensity have been shown to be insufficient for improving treatment outcome^40,41^. Instead, an individualized, function-oriented approach emphasizing daily activities, coping strategies, and quality of life has been recognized as essential for effective pain management ^42^. In this regards, studies highlighted the importance of developing (active) coping strategies—as the thoughts and actions in which people engage in their efforts to manage pain on a daily basis irrespective of the presence of pain—to better live with chronic pain through improved functionality ^43,44^. Enhancing physical functionality and coping mechanisms can facilitate return to daily activities and employment, reducing work absenteeism and societal burden, as CLBP remains a leading cause of disability and economic cost in industrialized countries ^45,46^. Overall, our findings underscore the value of a network-based perspective in understanding CLBP, illustrating how targeted interventions that enhance functionality and reduce pain interference may lead to broad improvements in psychological well-being and quality of life.

Based on our directed model, targeting fear of movement or catastrophizing alone—without interventions aimed at improving physical function—is likely suboptimal. Indeed, both fear of movement and catastrophizing were primarily receivers of causal influence, largely shaped by pain-related features. This aligns with reviews showing that while fear-avoidance beliefs predict outcomes in subacute low back pain, they have limited prognostic value in chronic pain contexts, although no causal effects were investigated ^47^. Using this finding along with what we found in our directed network— pain-related features influence fear of movement and catastrophizing— it can be speculated that fear and catastrophizing may be modified if participants are able to function without excessive pain or harm and targeting fear without movement-based interventions aiming improving function is not the best strategy. This interpretation supports integrative movement-based approaches such as graded exposure^48^, cognitive functional therapy ^49^, and/or pain reprocessing therapy ^50^ which combine physical engagement with cognitive and emotional retraining. Conversely, purely cognitive or psychological interventions tend to yield limited improvements in functional outcomes ^51^.

### 4.1 Strengths and limitations

The main strengths of this study include the large clinical sample of CLBP patients, who reported on a set of biopsychosocial features using the QLBPS dataset, enhancing the generalizability of findings to patients in Quebec and potentially Canada. Another strength lies in the direct comparison of analytical frameworks—mutual information, distance correlation, and LiNGAM— which allowed identification of the most suitable model for examining interrelations among biopsychosocial features and ensured replicability of results across methods. Furthermore, the integration of influence maximization with directed network analysis provided complementary insights into causal pathways and influential hubs within the system. Robustness checks further strengthened the findings, as bootstrapped confidence intervals showed stable accuracy of the strongest edge weights, and centrality stability analyses showed that central symptom rankings remained consistent until approximately 50% of the sample was dropped. The main limitation is the self-selection nature of the cohort. As the presence of LBP was based solely on self-report, we were unable to differentiate between specific and non-specific LBP, potentially introducing heterogeneity. Furthermore, psychosocial factors (e.g., depressive symptoms, sleep disturbance, fear of movement, catastrophizing) were assessed using single-item categorical measures from the NIH minimal dataset rather than validated multi-item questionnaires, which may have constrained measurement precision and influenced the network estimation.

## 5. Conclusion

This study provides a comprehensive network-based understanding of the complex interactions among biopsychosocial indicator in CLBP. Using complementary undirected and directed modeling approaches, we identified pain interference, physical function, and depression as the most central and influential indicator shaping the overall system. The LiNGAM analysis further revealed that physical function, depression, and pain interference exert causal effects on downstream variables such as fear of movement, catastrophizing, and widespread pain, while influence maximization and diffusion simulations confirmed their pivotal roles as drivers of network-wide activation. These findings underscore that interventions targeting functionality and pain interference—rather than pain intensity alone—may yield broader improvements across psychological and functional domains. Adopting a network-informed, multidimensional assessment framework can thus enhance individualized pain management aims at restoring daily functioning and quality of life in individuals with CLBP.

## Data Availability

All data produced in the present study are available upon reasonable request to the authors

## References

1. Newman M. Networks: An Introduction. Oxford University Press; 2010.

2. McNally RJ. The network takeover reaches psychopathology. Behavioral and Brain Sciences. 2019;42

3. Borsboom D, Cramer AO. Network analysis: an integrative approach to the structure of psychopathology. Annual review of clinical psychology. 2013;9(1):91–121.

4. Hartvigsen J, Hancock MJ, Kongsted A, et al. What low back pain is and why we need to pay attention. The Lancet. 2018;391(10137):2356–2367.

5. Costa LdCM, Maher CG, McAuley JH, et al. Prognosis for patients with chronic low back pain: inception cohort study. Bmj. 2009;339

6. Tanguay-Sabourin C, Fillingim M, Guglietti GV, et al. A prognostic risk score for development and spread of chronic pain. Nature Medicine. 2023;29(7):1821–1831.

7. Chou R, Shekelle P. Will this patient develop persistent disabling low back pain? Jama. 2010;303(13):1295–1302.

8. Stevans JM, Delitto A, Khoja SS, et al. Risk factors associated with transition from acute to chronic low back pain in US patients seeking primary care. JAMA network open. 2021;4(2):e2037371–e2037371.

9. Shmagel A, Foley R, Ibrahim H. Epidemiology of chronic low back pain in US adults: data from the 2009–2010 National Health and Nutrition Examination Survey. Arthritis care & research. 2016;68(11):1688–1694.

10. McWilliams LA, Sarty G, Kowal J, Wilson KG. A network analysis of depressive symptoms in individuals seeking treatment for chronic pain. The Clinical journal of pain. 2017;33(10):899–904.

11. Zhao X, Boersma K, Gerdle B, Molander P, Hesser H. Fear network and pain extent: Interplays among psychological constructs related to the fear-avoidance model. Journal of Psychosomatic Research. 2023;167:111176.

12. Penedo JMG, Rubel JA, Blättler L, et al. The complex interplay of pain, depression, and anxiety symptoms in patients with chronic pain: a network approach. The Clinical Journal of Pain. 2020;36(4):249–259.

13. Åkerblom S, Cervin M, Perrin S, Rivano Fischer M, Gerdle B, McCracken LM. A network analysis of clinical variables in chronic pain: a study from the Swedish quality registry for pain rehabilitation (SQRP). Pain Medicine. 2021;22(7):1591–1602.

14. Wi D, Park C, Ransom JC, Flynn DM, Doorenbos AZ. A network analysis of pain intensity and pain-related measures of physical, emotional, and social functioning in US military service members with chronic pain. Pain Medicine. 2024;25(3):231–238.

15. Pagé GM, Lacasse A, Beaudet N, et al. The Quebec Low Back Pain Study: A protocol for an innovative 2-tier provincial cohort. Pain Reports. 2020;5(1):e799.

16. Liu Z-Q, Luppi AI, Hansen JY, et al. Benchmarking methods for mapping functional connectivity in the brain. Nature Methods. 2025:1–10.

17. Shimizu S, Inazumi T, Sogawa Y, et al. DirectLiNGAM: A direct method for learning a linear non-Gaussian structural equation model. Journal of Machine Learning Research-JMLR. 2011;12(Apr):1225–1248.

18. Shimizu S, Hoyer PO, Hyvärinen A, Kerminen A, Jordan M. A linear non-Gaussian acyclic model for causal discovery. Journal of Machine Learning Research. 2006;7(10)

19. Page GM, Lacasse A, Quebec Back Pain C, et al. The Quebec Low Back Pain Study: a protocol for an innovative 2-tier provincial cohort. Pain Rep. Jan-Feb 2020;5(1):e799. doi:10.1097/PR9.0000000000000799

20. Masse-Alarie H, Angarita-Fonseca A, Lacasse A, et al. Low back pain definitions: effect on patient inclusion and clinical profiles. Pain Rep. Mar-Apr 2022;7(2):e997. doi:10.1097/PR9.0000000000000997

21. Angarita-Fonseca A, Pagé MG, Meloto CB, et al. The Canadian version of the National Institutes of Health minimum dataset for chronic low back pain research: reference values from the Quebec Low Back Pain Study. Pain. Feb 1 2023;164(2):325–335. doi:10.1097/j.pain.0000000000002703

22. von Elm E, Altman DG, Egger M, Pocock SJ, Gøtzsche PC, Vandenbroucke JP. The Strengthening the Reporting of Observational Studies in Epidemiology (STROBE) statement: guidelines for reporting observational studies. J Clin Epidemiol. Apr 2008;61(4):344–9. doi:10.1016/j.jclinepi.2007.11.008

23. Dionne CE, Dunn KM, Croft PR, et al. A consensus approach toward the standardization of back pain definitions for use in prevalence studies. Spine (Phila Pa 1976). Jan 1 2008;33(1):95–103. doi:10.1097/BRS.0b013e31815e7f94

24. Deyo RA, Dworkin SF, Amtmann D, et al. Report of the NIH Task Force on research standards for chronic low back pain. Physical therapy. 2015;95(2):e1–e18.

25. Harris PA, Taylor R, Thielke R, Payne J, Gonzalez N, Conde JG. Research electronic data capture (REDCap)--a metadata-driven methodology and workflow process for providing translational research informatics support. J Biomed Inform. Apr 2009;42(2):377–81. doi:10.1016/j.jbi.2008.08.010

26. Deyo RA, Dworkin SF, Amtmann D, et al. Focus article report of the NIH task force on research standards for chronic low back pain. The Clinical journal of pain. 2014;30(8):701–712.

27. Lacasse A, Roy JS, Parent AJ, et al. The Canadian minimum dataset for chronic low back pain research: a cross-cultural adaptation of the National Institutes of Health Task Force Research Standards. CMAJ Open. Jan-Mar 2017;5(1):E237–e248. doi:10.9778/cmajo.20160117

28. Pincus T, Burton AK, Vogel S, Field AP. A systematic review of psychological factors as predictors of chronicity/disability in prospective cohorts of low back pain. Spine. 2002;27(5):E109–E120.

29. Hruschak V, Cochran G. Psychosocial predictors in the transition from acute to chronic pain: a systematic review. Psychology, health & medicine. 2018;23(10):1151–1167.

30. Song L, Langfelder P, Horvath S. Comparison of co-expression measures: mutual information, correlation, and model based indices. BMC bioinformatics. 2012;13(1):328.

31. Hagberg A, Swart PJ, Schult DA. Exploring network structure, dynamics, and function using NetworkX. 2007.

32. Borgatti SP. Centrality and network flow. Social networks. 2005;27(1):55–71.

33. Epskamp S, Borsboom D, Fried EI. Estimating psychological networks and their accuracy: A tutorial paper. Behav Res Methods. Feb 2018;50(1):195–212. doi:10.3758/s13428-017-0862-1

34. Ikeuchi T, Ide M, Zeng Y, Maeda TN, Shimizu S. Python package for causal discovery based on LiNGAM. Journal of Machine Learning Research. 2023;24(14):1–8.

35. Wang C, Chen W, Wang Y. Scalable influence maximization for independent cascade model in large-scale social networks. Data Mining and Knowledge Discovery. 2012;25(3):545–576.

36. Åkerblom S, Cervin M, Perrin S, Rivano Fischer M, Gerdle B, McCracken LM. A Network Analysis of Clinical Variables in Chronic Pain: A Study from the Swedish Quality Registry for Pain Rehabilitation (SQRP). Pain Med. Jul 25 2021;22(7):1591–1602. doi:10.1093/pm/pnaa473

37. IsHak WW, Wen RY, Naghdechi L, et al. Pain and depression: a systematic review. Harvard review of psychiatry. 2018;26(6):352–363.

38. Martinez-Calderon J, Jensen MP, Morales-Asencio JM, Luque-Suarez A. Pain catastrophizing and function in individuals with chronic musculoskeletal pain: a systematic review and meta-analysis. The Clinical journal of pain. 2019;35(3):279–293.

39. Luque-Suarez A, Martinez-Calderon J, Falla D. Role of kinesiophobia on pain, disability and quality of life in people suffering from chronic musculoskeletal pain: a systematic review. British journal of sports medicine. 2019;53(9):554–559.

40. Scher C, Meador L, Van Cleave JH, Reid MC. Moving Beyond Pain as the Fifth Vital Sign and Patient Satisfaction Scores to Improve Pain Care in the 21st Century. Pain Management Nursing. 2018/04/01/ 2018;19(2):125–129.

41. Mularski RA, White-Chu F, Overbay D, Miller L, Asch SM, Ganzini L. Measuring pain as the 5th vital sign does not improve quality of pain management. J Gen Intern Med. Jun 2006;21(6):607–12. doi:10.1111/j.1525-1497.2006.00415.x

42. Mallick-Searle T, Sharma K, Toal P, Gutman A. Pain and Function in Chronic Musculoskeletal Pain-Treating the Whole Person. J Multidiscip Healthc. 2021;14:335–347. doi:10.2147/jmdh.S288401

43. Zeidner M, Endler NS. Handbook of coping: Theory, research, applications. John Wiley & Sons; 1995.

44. Van Damme S, Crombez G, Eccleston C. Coping with pain: a motivational perspective. Pain. Sep 30 2008;139(1):1–4. doi:10.1016/j.pain.2008.07.022

45. Hoy D, March L, Brooks P, et al. The global burden of low back pain: estimates from the Global Burden of Disease 2010 study. Annals of the rheumatic diseases. 2014;73(6):968–974.

46. Maher C, Underwood M, Buchbinder R. Non-specific low back pain. The Lancet. 2017;389(10070):736–747.

47. Wertli MM, Rasmussen-Barr E, Weiser S, Bachmann LM, Brunner F. The role of fear avoidance beliefs as a prognostic factor for outcome in patients with nonspecific low back pain: a systematic review. Spine J. May 1 2014;14(5):816–36.e4. doi:10.1016/j.spinee.2013.09.036

48. Macedo LG, Smeets RJ, Maher CG, Latimer J, McAuley JH. Graded activity and graded exposure for persistent nonspecific low back pain: a systematic review. Phys Ther. Jun 2010;90(6):860–79. doi:10.2522/ptj.20090303

49. O’sullivan P. It’s time for change with the management of non-specific chronic low back pain. BMJ Publishing Group Ltd and British Association of Sport and Exercise Medicine; 2012. p. 224–227.

50. Ashar YK, Gordon A, Schubiner H, et al. Effect of Pain Reprocessing Therapy vs Placebo and Usual Care for Patients With Chronic Back Pain: A Randomized Clinical Trial. JAMA Psychiatry. Jan 1 2022;79(1):13–23. doi:10.1001/jamapsychiatry.2021.2669

51. Williams ACdC, Fisher E, Hearn L, Eccleston C. Psychological therapies for the management of chronic pain (excluding headache) in adults. Cochrane Database of Systematic Reviews. 2020;(8)doi:10.1002/14651858.CD007407.

