## supplemental file for "Development of a network for interactions and associations among biopsychosocial features of chronic low back pain"

**eResults**

**eTable 1.** Selected features used for creating networks along with their measurements in from Canadian adaptation of NIH minimal dataset

| **Order** | **Features** | **Labels in networks** | **Measurement** |
| --- | --- | --- | --- |
| **1** | Age | age | Date of Birth  YYYY-MM-DD |
| **2** | Sex | sex | Sex (at birth)   - Female - Male - Unknown - Unspecified |
| **3** | Employment status | emplo | Employment status   - Working now fulltime - Working now part-time - Looking for work, unemployed - Sick leave or maternity leave - Disabled due to back pain, permanently or temporarily - Disabled for reasons other than back pain - Student - Temporarily laid off - Retired - Keeping house - Unknown - Other |
| **4** | Educational level | edu | Education level   - No high school diploma - High school (secondary school) diploma or equivalent - Registered apprenticeship or other trade certificate or diploma (e.g., hairstyling, cooking, electrician, carpentry, etc.) - College, CEGEP or other non-university certificate or diploma (e.g., accounting technology, industrial engineering technology, legal assistant, pre-university program, etc.) - University certificate or diploma below bachelor's level (e.g., undergraduate certificate) - Bachelor's degree (e.g., B.A., B.A. (Hons.), B.Sc., B.Ed., LL.B.) - University certificate or diploma above bachelor's level (e.g., D.E.S.S., Short Graduate Program) - Mater's degree (e.g., M.A., M.Sc., M.Ed., M.B.A.) - Degree in medicine, dentistry, veterinary medicine or optometry (M.D., D.D.S., D.M.D.,D.V.M., O.D.) - Doctorate (e.g., Ph.D., Psy.D, Ed.D.) |
| **5** | Quality of life | qol | Now, please write the number that indicates how your health is TODAY   - Min: 0 - Max: 100 |
| **6** | Sleep disturbance | sleep | 1. My sleep quality was 2. My sleep was refreshing 3. I had a problem with my sleep 4. I had difficulty falling asleep   Ratio for each question   - Not at all - A little bit - Somewhat - Quite a bit - Very much |
| **7** | Body mass index | bmi | Calculated using weight and height  weight (kg) / [height (m)]^2^ |
| **8** | Smoking habit | smok | How would you describe your cigarette smoking?   - Never smoked - Current smoker - Used to smoke, but have now quit |
| **9** | Comorbidity | comor | Do you have any of the following conditions?  Do you receive treatment for it?  Does it limit your activities?   1. Heart disease 2. High blood pressure 3. Lung disease 4. Diabetes 5. Ulcer or stomach disease 6. Kidney disease 7. Anaemia or other blood disease 8. Cancer 9. Depressive symptoms 10. Osteoarthritis, degenerative arthritis 11. Back pain 12. Rheumatoid arthritis 13. Other medical problems   Ratio for each question   - Yes= 1 - No= 0 |
| **10** | Pain intensity | pain | In the past 7 days, how would you rate your low back pain on average?   - 0 = No pain - 10 = Worst imaginable pain |
| **11** | Pain duration | pdur | How long has low back pain been an ongoing problem for you?   - Less than a month - 1-2 months - 3-5 months - 6-11 months - 1-5 years - More than 5 years |
| **12** | Pain interference | pinterf | 1. How much did pain interfere with your day-to-day activities? 2. How much did pain interfere with work around the home? 3. How much did pain interfere with your ability to participate in social activities? 4. How much did pain interfere with your household chores?   Ratio for each question   - Not at all - A little bit - Somewhat - Quite a bit - Very much |
| **13** | Widespread pain | widsp | Widespread pain (pain in most of your body)   - Not bothered at all - Bothered a little - Bothered a lot |
| **14** | Neuropathic pain | DN4 | Is the pain associated with one or more of the following symptoms in the same area?   1. Tingling 2. Pins and needles 3. Numbness 4. Itching   Ratio for each question   - Yes - No |
| **15** | Physical function | phyfunc | 1. Are you able to do chores such as vacuuming or yard work? 2. Are you able to go up and down stairs at a normal pace? 3. Are you able to go for a walk of at least 15 minutes? 4. Are you able to run errands and shop?   Ratio for each question   - Without any difficulty - With a little difficulty - With some difficulty - With much difficulty - Unable to do |
| **16** | Pain impact startification | impcat | Nine questions including the pain intensity, 4 items of pain interference and 4 items of physical function  From 8 (least impact) to 50 (most impact) and classified as   - Mild (8-27 points) - Moderate (28-34 points) - Severe (35-50 points) |
| **17** | Fear of movement | fear | It's not really safe for a person with my low back problem to be physically active   - Agree - Disagree |
| **18** | Catastrophizing | catstro | I feel that my low back pain is terrible and it's never going to get any better   - Agree - Disagree |
| **19** | Depressive symptoms | depp | 1. I felt worthless 2. I felt helpless 3. I felt depressed 4. I felt hopeless   Ratio for each question   - Never - Rarely - Sometimes - Often - Always |

- **Features’ collinearity**

Based on the efigure 1 the pain impact feature was strongly correlated (r ≥ 0.80) with pain interference and physical function. As pain impact includes four questions of pain interference and physical function and it replicates other two features, we removed it from the analysis to avoid multicollinearity issues.

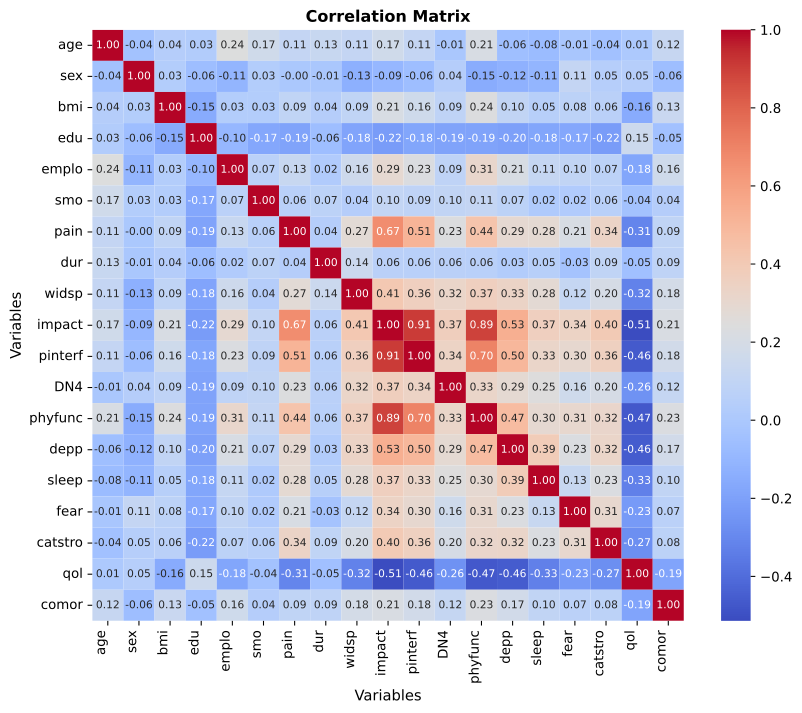

**eFigure 1.** Heatmap of pairwise distance correlations between all features. Warmer colors indicate stronger positive associations, cooler colors indicate negative associations. The pain impact feature was removed due to collinearity with pain interference and physical function (r ≥ 0.80).

- **Undirected network via mutual information and comparison of centrality with distance correlation model**

eFigure 2A presents the undirected network constructed using mutual information (MI). Overall, the MI-based network revealed patterns largely consistent with the distance correlation (dCor) model. In both networks, the strongest and most stable edges connected physical function, pain interference, and depressive symptoms, indicating these features form the central core of the biopsychosocial system. Additionally, pain intensity and quality of life showed strong and dense associations with the network core, reinforcing their integrative roles across domains. Conversely, features such as sex, smoking habit, and pain duration occupied more peripheral positions, characterized by weaker connections with the rest of the system, mirroring the pattern observed in the dCor model. Notably, however, the MI network differed in that catastrophizing and widespread pain were positioned further from the central cluster than in the dCor model, suggesting reduced connectivity and influence under MI-based estimation.

The Independent Cascade (IC) diffusion network based on MI is presented in Figure 2B. In this model, diffusion originated mainly and solely from physical function, identifying it as the most influential node within the MI network. Compared to dCor model, both models identified a single primary diffusion hub, but the spread and gradient of activation differed markedly. In the dCor model, diffusion initiated from pain interference, with secondary activation observed in physical function, and depressive symptoms, as indicated by the gradual color gradient extending across multiple nodes. This pattern reflects a more distributed influence consistent with widespread nonlinear dependencies among these features. In contrast, the MI-based diffusion was more localized, with physical function acting as a dominant but narrowly focused seed. Activation intensity declined sharply across the remaining nodes, as shown by the rapid color fade from the seed outward. This suggests that, under MI estimation, network influence is concentrated around physical function, with substantially weaker transmission across distal nodes. Together, these results indicate that while both models converge on physical function and pain interference as key diffusion hubs, the dCor network captures a broader and more graded propagation pattern, implying stronger integration among pain-related and psychological domains.

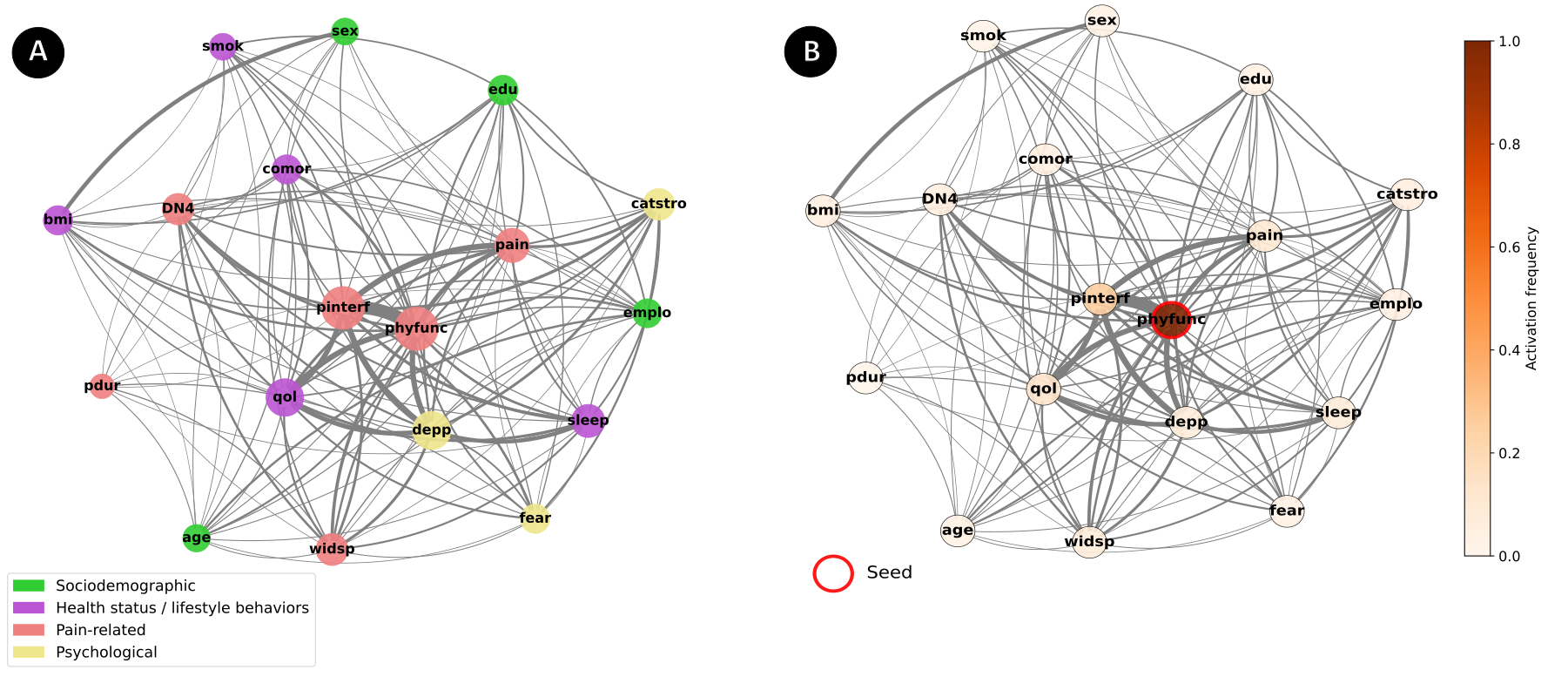

**eFigure 2.** Mutual information networks of socio-demographic, pain-related, and psychological features. **A:** Undirected network including all features and edges. Nodes represent individual variables (green = sociodemographic, purple = health status/lifestyle behaviors, red = pain-related, yellow = psychological). Edges represented by gray color showed pairwise distance correlations; edge thickness is proportional to correlation strength. Node size reflects node strength (sum of absolute edge weights). B: Independent Cascade diffusion in the MI undirected network. The figure illustrates the simulated diffusion process using the IC model, in which activation initiated from physical function (highlighted by a red ring) spreads probabilistically through the network according to edge weights. Node color intensity represents the activation frequency with darker shades indicating a higher probability of being activated during the diffusion process. Edges are scaled by their connection strength (weight magnitude) derived from MI coefficients. The simulation was repeated 1000 times to ensure stability of diffusion estimates, and node colors represent the average activation frequency across runs (darker shades = higher probability of activation). A base activation probability of 0.5 was used, meaning each active node had a 50% chance of activating a connected neighbor, scaled by the strength of their pairwise association (edge weight). Abbreviations: age= age; sex= sex; emplo = employment status; edu= educational level; qol = quality of life; sleep = sleep disturbance; bmi= Body mass index; smok= smoking habit; comor = comorbidity; pain = pain intensity; pdur= pain duration; pinterf = pain interference; widsp = widespread pain; DN4 = neuropathic pain; phyfunc = physical function; fear = fear of movement; catstro = catastrophizing; depp = depressive symptoms

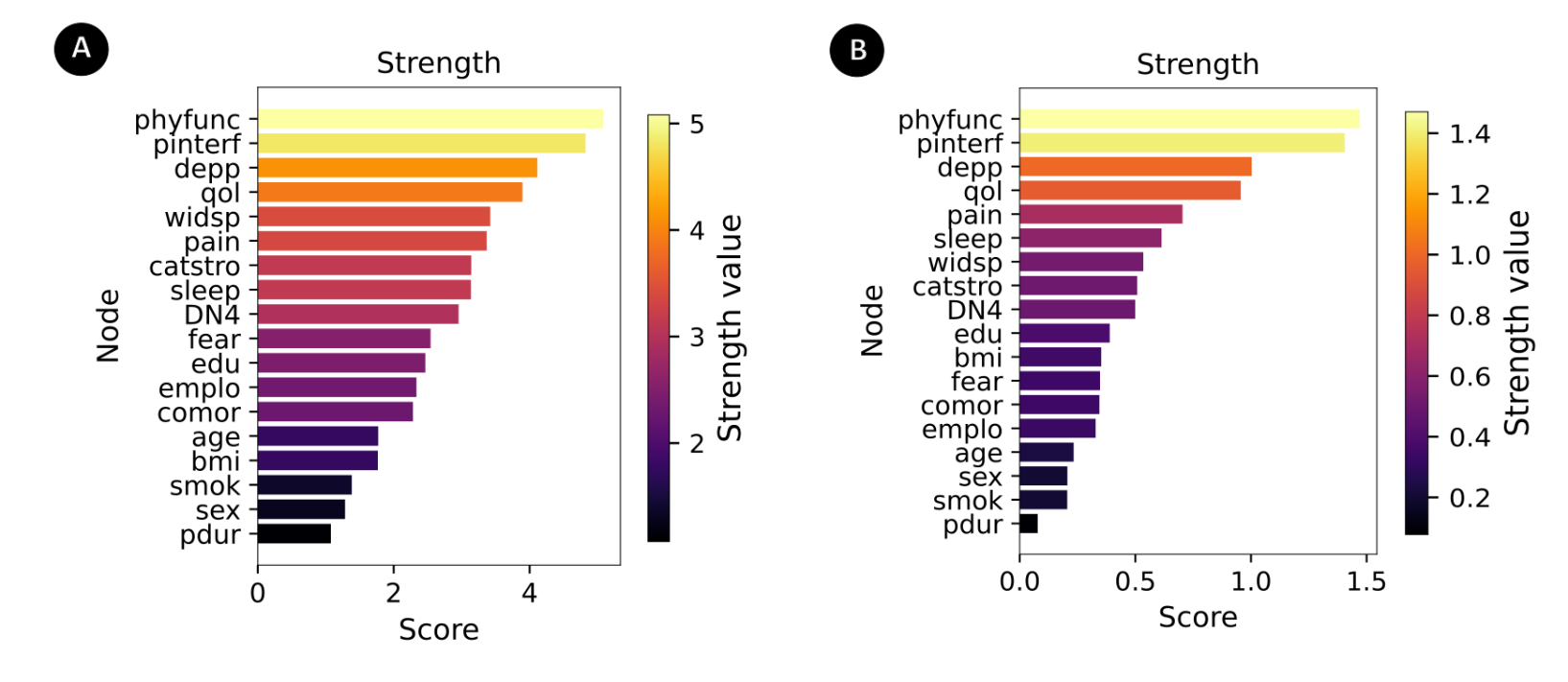
Ranking of strength centrality metric among selected features in distance correlation (dCore) and mutual information (MI) models are represented in eFigure 3A and 3B, respectivly. Strength is defined as the sum of all edge weights connected to a node ^1^. This measure reflects each node’s topological prominence, with higher strength indicating stronger integration within the network ^1^. eFigure 3 compares the strength centrality distributions derived from the dCor (panel A) and MI panel B) networks. Both models identified the same highest-valued strength centrality nodes—physical function, pain interference, depressive symptoms, and quality of life—and the same lowest-valued nodes, including sex, smoking, and pain duration.

**eFigure 3.** Strength centrality in undirected networks derived from dCor and MI. **A:** Strength centrality values from the dCor network. **B:** Strength centrality values from the MI network. Bar length and color intensity indicate the normalized strength centrality, representing the sum of absolute edge weights connected to each node. Both models identified physical function, pain interference, depressive symptoms, and quality of life as nodes with the highest strength centrality, whereas sex, smoking, and pain duration exhibited the lowest values. Abbreviations: age= age; sex= sex; emplo = employment status; edu= educational level; qol = quality of life; sleep = sleep disturbance; bmi= Body mass index; smok= smoking habit; comor = comorbidity; pain = pain intensity; pdur= pain duration; pinterf = pain interference; widsp = widespread pain; DN4 = neuropathic pain; phyfunc = physical function; fear = fear of movement; catstro = catastrophizing; depp = depressive symptoms

Examining the accuracy and stability of our network via bootstrapping with number of bootstrap samples of 1000, we found the strongest and most stable edge was between physical function and pain interference (edge weight= 0.68, 95% CI: 0.67–0.70). Additional robust associations included depressive symptoms with pain interference (edge weight= 0.48, 95% CI: 0.46–0.50), pain intensity with pain interference (edge weight= 0.47, 95% CI: 0.45–0.49), and depressive symptoms with physical function (edge weight= 0.45, 95% CI: 0.43–0.47). A somewhat weaker but still reliable connection was observed between physical function and quality of life (edge weight ≈ 0.44, 95% CI: 0.42–0.45). Overall, the bootstrap results highlight a dense core of strong and precise associations centered on pain interference and physical function, with depressive symptoms further bridging to both pain-related and quality-of-life domains (eFigure 4A).

Centrality stability analysis indicated a correlation stability coefficient of 0.50, meaning that up to 50% of the sample could be removed while centrality ranks still correlated at τ ≥ 0.70 with 95% probability. Stability curves further supported this result: median Kendall’s τ values gradually declined as larger proportions of cases were dropped, yet strength centrality remained above the τ = 0.70 threshold until approximately half of the data were removed (eFigure 4B). These findings suggest that centrality estimate from the dCor network is moderately to highly stable, allowing reliable interpretation of node importance, though caution is warranted when differentiating between nodes with very similar ranks.

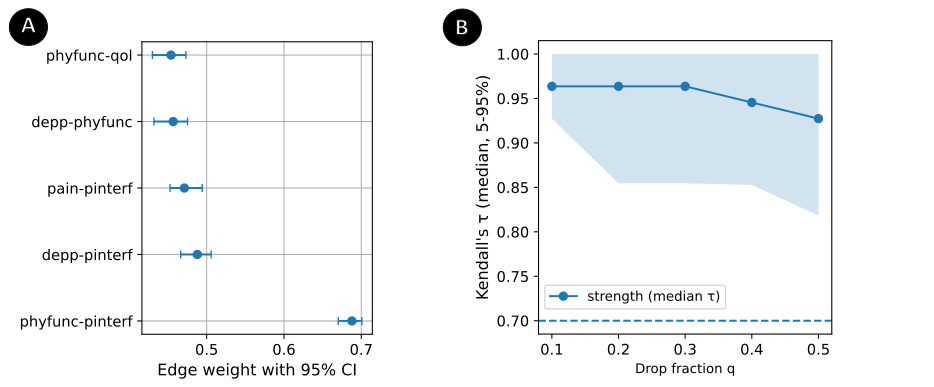

**eFigure 4.** Edge weight and centrality stability of the distance correlation (dCor) network. **A:** Bootstrap confidence intervals (95% CI) for the strongest network edges. Edges between physical function-pain interference, depressive symptoms -pain interference, pain intensity-pain interference, depressive symptoms -physical function, and physical function- quality of life showed the top five robust associations, with consistently high edge weights across bootstrap samples. **B:** Rank stability analysis for centrality metric of strength. The solid line represents the median Kendall’s τ correlation (τ_q50) between centrality ranks in the network and those in subsampled networks after dropping a fraction q of cases. Shaded area indicates the variability (5th–95th percentile range, τ_q05–τ_q95). The dashed horizontal line marks τ = 0.70, a commonly used threshold for acceptable stability. Values above this line suggest that centrality ranking remain stable under subsampling, whereas values approaching the threshold indicate reduced reliability. Abbreviations: Abbreviations: age= age; sex= sex; emplo = employment status; edu= educational level; qol = quality of life; sleep = sleep disturbance; bmi= Body mass index; smok= smoking habit; comor = comorbidity; pain = pain intensity; pdur= pain duration; pinterf = pain interference; widsp = widespread pain; DN4 = neuropathic pain; phyfunc = physical function; fear = fear of movement; catstro = catastrophizing; depp = depressive symptoms

- **Directed network**

**eTable 2.** Features according to their weight values of causal effects, ranked by weight

| **Source** | **Target** | **Weight** |
| --- | --- | --- |
| Physical function | Pain interference | 0.45 |
| Pain intensity | Catastrophizing | 0.45 |
| Neuropathic pain | Widespread pain | 0.37 |
| Catastrophizing | Fear of movement | 0.34 |
| Pain interference | Pain intensity | 0.28 |
| Depressive symptoms | Catastrophizing | 0.27 |
| Widespread pain | Catastrophizing | 0.25 |
| Age | Employment status | 0.24 |
| Pain interference | Depressive symptoms | 0.24 |
| Employment status | Fear of movement | 0.24 |
| Sleep disturbance | Depressive symptoms | 0.21 |
| Pain interference | Fear of movement | 0.21 |
| Neuropathic pain | Fear of movement | 0.20 |
| Age | Smoking habit | 0.19 |
| Physical function | Fear of movement | 0.19 |
| DN4 | Catastrophizing | 0.17 |
| Depressive symptoms | Widespread pain | 0.16 |
| Depressive symptoms | Fear of movement | 0.16 |
| Pain duration | Catastrophizing | 0.15 |
| Pain interference | Catastrophizing | 0.15 |
| Pain intensity | Widespread pain | 0.15 |
| Pain intensity | Fear of movement | 0.15 |
| Sleep disturbance | Pain intensity | 0.14 |
| Pain duration | Widespread pain | 0.13 |
| Age | Widespread pain | 0.13 |
| Sleep disturbance | Widespread pain | 0.12 |
| Sex | Fear of movement | 0.12 |
| Depressive symptoms | Employment status | 0.12 |
| Physical function | BMI | 0.11 |
| Pain interference | Neuropathic pain | 0.11 |
| Depressive symptoms | Neuropathic pain | 0.11 |
| Widespread pain | fear | 0.10 |
| Comorbidity | Widespread pain | 0.10 |
| Physical function | Employment status | 0.10 |
| Widespread pain | Employment status | 0.10 |
| BMI | Widespread pain | 0.10 |
| Pain interference | Widespread pain | 0.09 |
| Pain intensity | Neuropathic pain | 0.09 |
| Comorbidity | Employment status | 0.09 |
| Comorbidity | BMI | 0.08 |
| Physical function | Depressive symptoms | 0.08 |
| Sleep disturbance | Neuropathic pain | 0.08 |
| Comorbidity | Physical function | 0.08 |
| Smoking habit | Catastrophizing | 0.08 |
| Depressive symptoms | Pain intensity | 0.07 |
| Age | Educational level | 0.07 |
| Neuropathic pain | Smoking habit | 0.07 |
| Pain interference | Employment status | 0.06 |
| Comorbidity | Pain duration | 0.06 |
| Physical function | Catastrophizing | 0.06 |
| Physical function | Neuropathic pain | 0.06 |
| Physical function | Pain intensity | 0.06 |
| Comorbidity | Depressive symptoms | 0.06 |
| Pain interference | BMI | 0.05 |
| Smoking habit | Employment status | 0.05 |
| Depressive symptoms | Smoking habit | 0.04 |
| Physical function | Widespread pain | 0.04 |
| Pain duration | Smoking habit | 0.04 |
| Catastrophizing | Employment status | 0.02 |
| Age | Neuropathic pain | -0.05 |
| Depressive symptoms | Educational level | -0.05 |
| Educational level | Employment status | -0.06 |
| Sleep disturbance | Educational level | -0.07 |
| Comorbidity | Qualiy of life | -0.07 |
| Pain intensity | Educational level | -0.07 |
| Neuropathic pain | Educational level | -0.10 |
| Qualiy of life | BMI | -0.11 |
| Physical function | Qualiy of life | -0.11 |
| Sleep disturbance | Qualiy of life | -0.13 |
| Qualiy of life | Employment status | -0.13 |
| Qualiy of life | Neuropathic pain | -0.13 |
| Age | Catastrophizing | -0.13 |
| Pain duration | Fear of movement | -0.14 |
| BMI | Educational level | -0.15 |
| Pain interference | Qualiy of life | -0.16 |
| Educational level | Smoking habit | -0.16 |
| Pain intensity | Qualiy of life | -0.17 |
| Educational level | Widespread pain | -0.17 |
| Educational level | Fear of movement | -0.22 |
| Qualiy of life | Fear of movement | -0.27 |
| Qualiy of life | Widespread pain | -0.28 |
| Depressive symptoms | Qualiy of life | -0.28 |
| Qualiy of life | Catastrophizing | -0.31 |
| Educational level | Catastrophizing | -0.31 |

**eTable 3.** Features according to their out-degree and in-degree centrality values, ranked by out-degree

| **Features** | **Out_Degree (influencer)** | **In_Degree (influence received)** |
| --- | --- | --- |
| Physical function | 10 | 1 |
| Depressive symptoms | 9 | 4 |
| Pain interference | 9 | 1 |
| Comorbidity | 7 | 0 |
| Pain intensity | 6 | 0 |
| Sleep disturbance | 6 | 6 |
| Qualiy of life | 6 | 4 |
| Age | 6 | 0 |
| Neuropathic pain | 5 | 7 |
| Educational level | 5 | 6 |
| Pain duration | 4 | 1 |
| Widespread pain | 3 | 12 |
| Smoking habit | 3 | 5 |
| Catastrophizing | 2 | 11 |
| Employment status | 2 | 10 |
| BMI | 2 | 4 |
| Sex | 1 | 0 |
| Fear of movement | 0 | 12 |

Bibliography

1.	Borgatti SP. Centrality and network flow. *Social networks*. 2005;27(1):55-71.
